# Risk of aortic aneurysm in patients with psoriasis: a systematic review and meta-analysis

**DOI:** 10.1101/2020.04.03.20052191

**Authors:** Xinyu Yu, Xin Feng, Liangtao Xia, Shiyi Cao, Xiang Wei

## Abstract

**Objective:** Psoriasis is a chronic inflammatory disease that may play a role in the pathogenesis of atherosclerotic vascular diseases. We conducted this systematic review to investigate the association between psoriasis and the risk of aortic aneurysm.

**Methods:** Pubmed, Embase, and Scopus were searched up to July 20, 2019. We included cohort studies if they reported estimate effects assessing the association between having psoriasis and aortic aneurysm. We used Newcastle-Ottawa Scale to evaluate methodology quality of eligible studies. Random-effect meta-analyses were used to pool individual estimates. Subgroup analyses were conducted to find heterogeneity source.

**Results:** After a view of 2207 citations, we included three large cohort studies enrolling 5,706,525 participants in this systematic review. Psoriasis patients have an increased risk of development of aortic aneurysm (hazard ratio [HR]: 1.30, 95%confidence intervals [CI], 1.10 to 1.55, *I*^2^ = 53.1%). However, the risk of AA was not significantly increased in female psoriasis patients (HR, 1.55, 95%CI, 0.65 to 3.72), patients with hypertension (HR, 1.44, 95%CI, 0.85 to 2.42), patients with hyperlipidemia (HR, 1.69, 95%CI, 1.15 to 2.48) and patients with diabetes (HR, 1.15, 95%CI 0.46 to 2.87).

**Conclusions:** Current evidence from observational studies suggests that psoriasis may increase the risk of aortic aneurysm, and screening of aortic aneurysm might be considered among psoriasis patients.

**Key questions:**

1. What is already known on this subject? Patients with psoriasis have an increased prevalence of cardiovascular diseases, including ischemic heart disease, heart failure, peripheral vascular disease and stroke. The increased risk of cardiovascular events is believed to be associated with the systematic inflammatory pathophysiological mechanisms of psoriasis.
2. What might this study add? This systematic review and meta-analysis summarized epidemiological evidence investigating the relationship between psoriasis and aortic aneurysm, and we found psoriasis patients have an increased risk of aortic aneurysm.
3. How might this impact on clinical practice? Screening for aortic aneurysm could be conducted among psoriasis patients, and the anti-inflammatory therapy may be helpful for the treatment of aortic aneurysm.

## 1. Introduction

Psoriasis is a systemic inflammatory disease that affects an estimated 3% of the US adult population[1]. Patients with psoriasis have an increased prevalence of cardiovascular diseases, including ischemic heart disease[2], heart failure[3], peripheral vascular disease[4] and stroke[5]. The increased risk of cardiovascular events is believed to be associated with the systematic inflammatory pathophysiological mechanisms of psoriasis[6].Moreover, aortic vascular inflammation detected by ^18^Fluorodeoxyglucose(FDG) positron emission tomography/ computed tomography (PET/CT) is associated with psoriasis skin disease severity[1, 7].

Aortic vascular inflammation plays an essential role in the development and progression of aortic aneurysm (AA). Chronic aortic vascular inflammation is believed to lead to destruction of the aortic media and to vascular smooth muscle cell dysfunction as a result of the release of a range of proteolytic enzymes, such as matrix metalloproteinases and cysteine proteases, oxidation-derived free radicals, cytokines, and related products[8]. Given these foundations, several studies[9, 10, 11] have explored the relative risk of AA in patients with psoriasis. However, these study findings are conflicting, and whether psoriasis increases the risk of AA is still unclear. Thus, we conducted this systematic review to investigate the association between psoriasis and the risk of AA.

## 2. Methods

### 2.1 Search strategy

The systematic review followed the Preferred Reporting Items for Systematic Reviews and Meta-analyses (PRISMA) guidelines. We conducted a systematic search on Pubmed, Scopus, and Embase for relevant full-text articles published before July 20, 2019. We did not set any language limitation in the literature search. Search terms (displayed in Electronic supplementary material, ESM) included a combination of keywords relating to psoriasis (e.g. ‘psoriatic arthritis’), aortic vascular inflammation (e.g. ‘vascular inflammation’) and aortic aneurysm (e.g. ‘aorta’, ‘aortic aneurysm’). To capture other potentially relevant articles, we manually checked the references of included literature.

### 2.2 Inclusion criteria and study selection

To clarify the association between psoriasis and the risk of AA, we included observational studies that satisfied the following criteria: studies reported effect estimates on the risk of aortic aneurysm in patients with psoriasis compared to healthy subjects; psoriasis patients undergoing phototherapy, topical therapy, oral-systemic medications or biologic agents for psoriasis treatment were eligible; aortic aneurysm of participants could be thoracic aortic aneurysm or abdominal aortic aneurysm. Conference abstracts were excluded as their full study reports could not be assessed and their scientific rigor had not been peer-reviewed[12]. Case reports and case series were excluded for lack of strict study design. Two investigators (XY and XF) independently screened identified articles to find eligible studies, and the senior investigator (SC) solved discrepancies in study selection.

### 2.3 Data extraction

We extracted the following information from eligible studies: (1) authors, (2) published year, (3) participants, (4) inclusion criteria of psoriasis patients, (5) definition of psoriasis severity, (6) diagnostic criteria of AA, (7) characteristics of control groups, (8) mean follow-up duration, (9) unadjusted and adjusted hazard ratios (HRs) with corresponding 95% confidence interval (CI), (10) adjustment of covariates, and (11) baseline characteristics of study population (age, gender and comorbidity).Detailed information was presented in tables.

### 2.4 Quality appraisal

We applied the Newcastle-Ottawa Scale (NOS) to appraise the methodological quality of included studies. The scale is a scoring system covering three perspectives of methodology: selection of study population; comparability; and ascertainment of outcome and exposure. We did not exclude any study on the basis of quality appraisal. Two investigators (XY and XF) individually assessed the quality of eligible studies, and a senior investigator(SC) solved the discrepancies.

### 2.5 Statistical analyses

Unadjusted and adjusted HRs with corresponding 95%CI were extracted from included studies. We preferred adjusted HRs to reduce the impact of confoundings on estimate effects, and unadjusted HRs were used if adjusted estimates were not available. Some covariates (e.g. smoking) were not analyzed in original studies, and we calculated estimates using original data to investigate the effect. We used random-effect model meta-analyses to evaluate the association between psoriasis and AA. Statistical heterogeneity was quantified by using the inconsistency index (*I*^2^) test[13]. *I*^2^ values ranged from 0% to 100% and I^2^<50% was considered as low heterogeneity, and I^2^ value 50% to 75% as moderate heterogeneity, and I^2^>75% as statistically high heterogeneity. When we analyzed the overall risk of AA in patients with psoriasis and the overall HR was not provided, we used fixed-effect meta-analysis to calculate overall HR from separate HR. Due to the limited number of included study, we did not conduct a sensitivity analysis. We performed subgroup analyses to investigate confounding factors (e.g. age, gender, and comorbidities) that may interact with the association of interest. We summarized baseline characteristics of individual study. Student’s t-test and Chi-square test were used to analyze the difference of characteristics in psoriasis patients and reference population. All P value were two-tailed and P<0.05 was set as the significance level. All analyses were conducted in Stata version 14.0.

### 2.6 Patient and Public Involvement statement

We conducted a systematic review of published studies, and patients involved in the included studies. We did not recruit other patients in this article.

## 3. Results

### 3.1 Study characteristics and quality appraisal

The searching strategy was displayed in Figure 1. In total, 2207 records were identified, and 8 articles were assessed for eligibility after screening. Finally, three studies[9, 10, 11] with 5,706,525 participants were included in this systematic review. The included studies were scored 7 to 8 points in NOS (see Supplementary Table 1 in ESM). Detailed characteristics of included studies and baseline characteristics of included population were arranged in Table 1 and Supplementary Table 2. Three studies were performed in USA, Denmark, and Taiwan respectively. Basic features (age, sex, and comorbidity) were significantly different between psoriasis cases and reference population, and we used HRs adjusted by these factors (see Table1) to avoid the interference of them on pooled results.

**Table 1.**
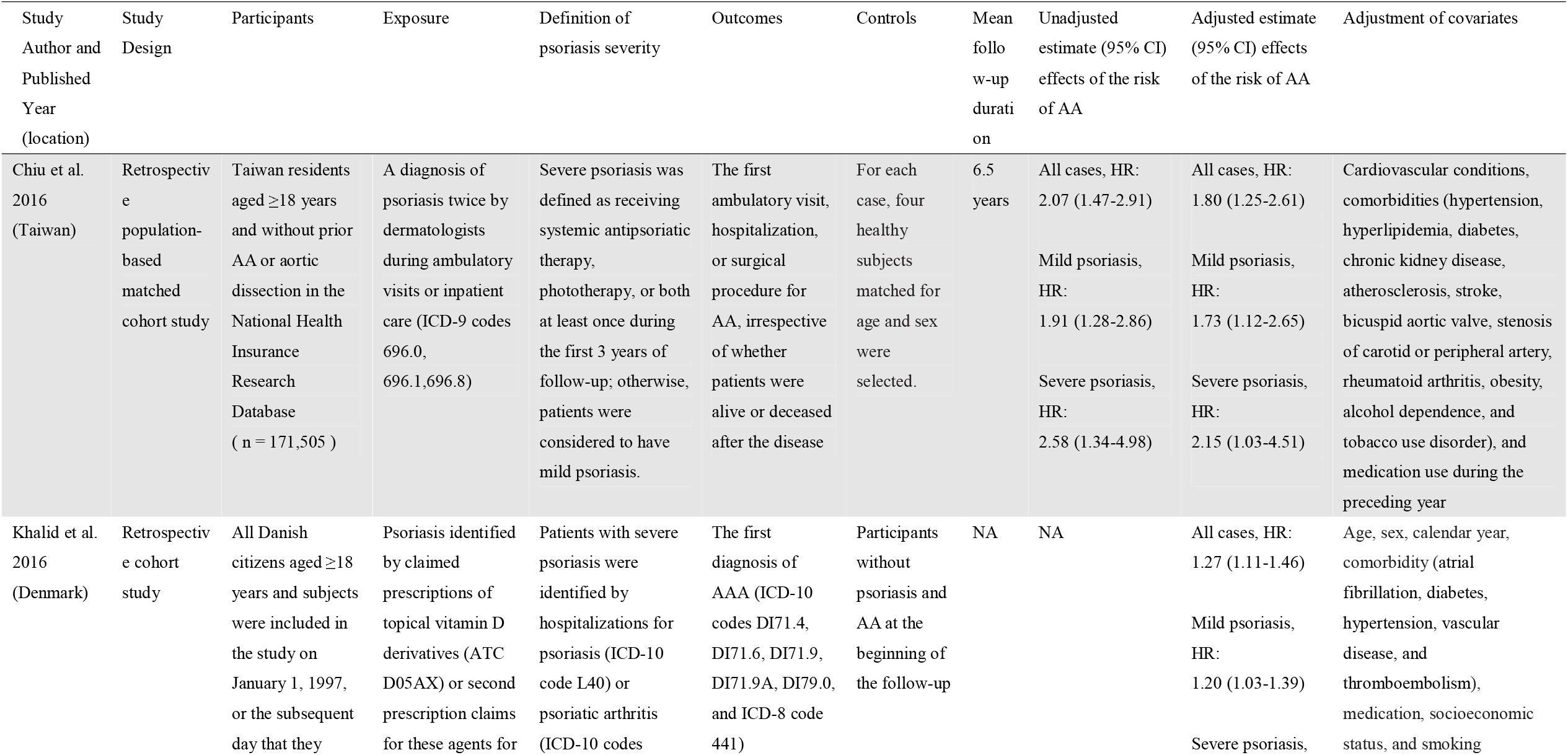
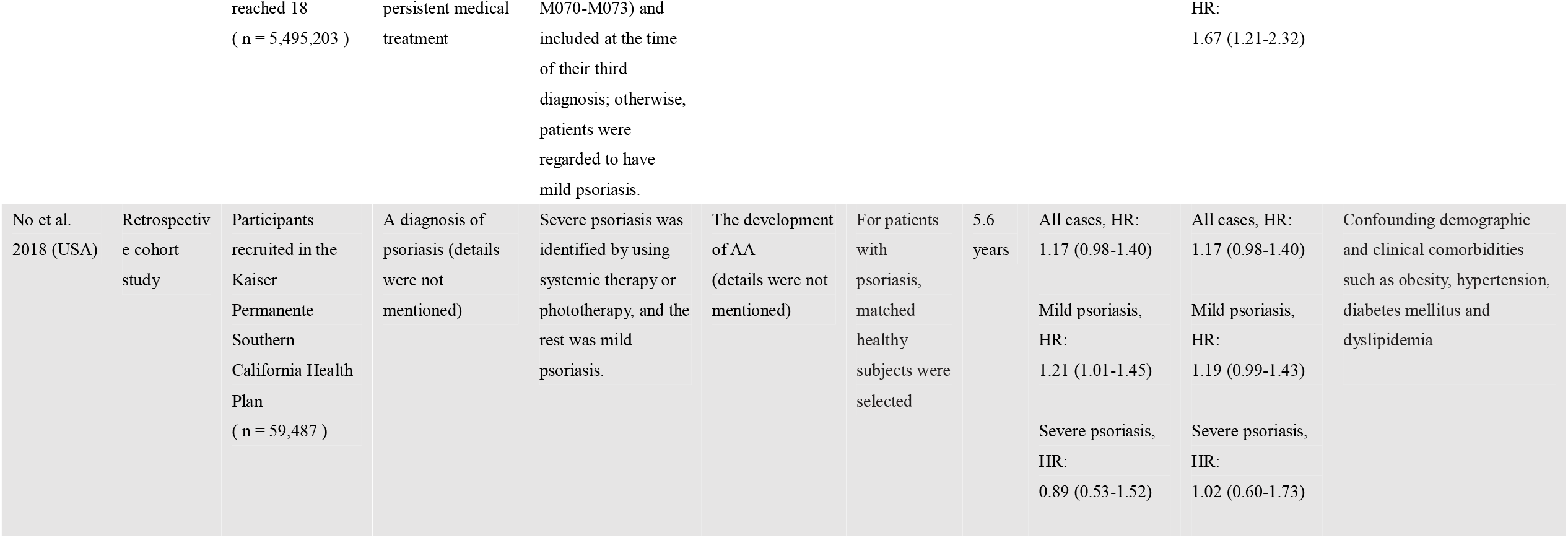
Characteristics of studies included in the meta-analysis.

**Figure 1.**
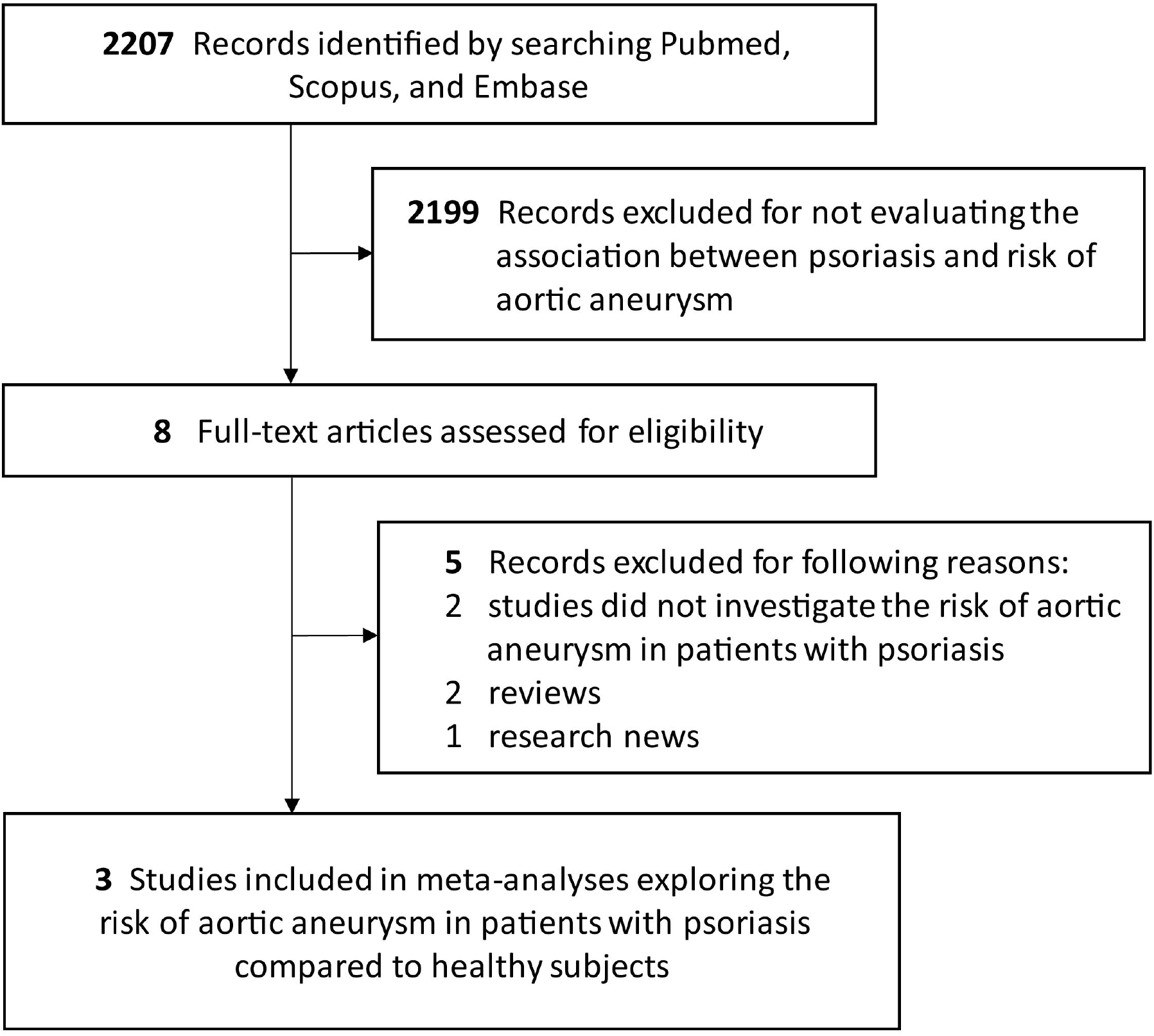
Study screening flowchart

### 3.2 Risk of AA in patients with psoriasis

Forest plots of meta-analyses were prepared in Figure 2. Three studies with 24 864 cases indicated an HR of 1.30 (95%CI, 1.10 to 1.55, *I*^2^ = 53.1%) for association between the risk of AA and psoriasis. Publication bias could not be effectively evaluated using 3 studies, and thus the result may be biased.

**Figure 2.**
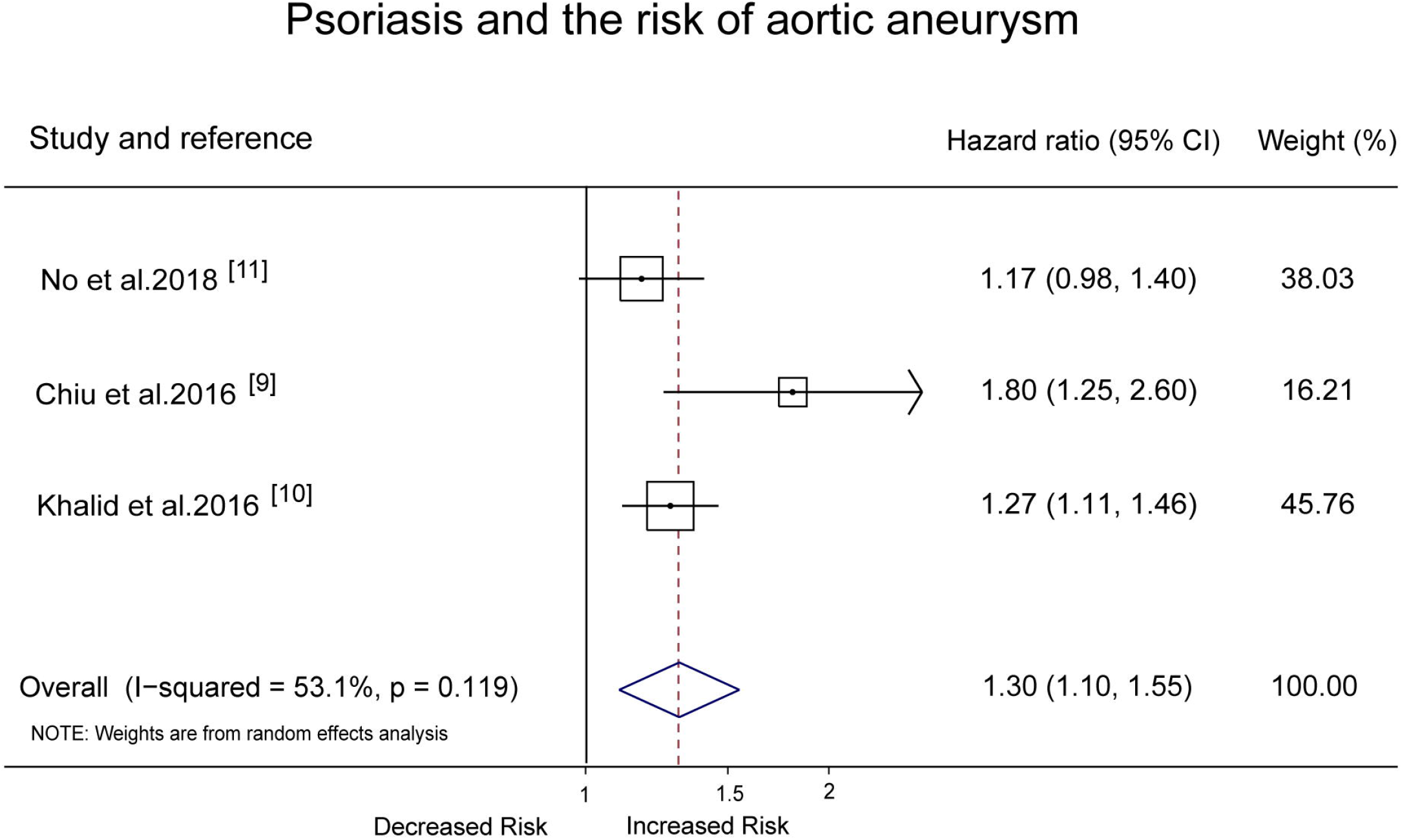
Forest plots of the meta-analyses for the risk of AA in patients with psoriasis Abbreviations: CI, confidence interval.

### 3.3 Subgroup analysis

According to the subgroup analysis (see Table 2), psoriasis could increase the risk of abdominal aortic aneurysm and thoracic aortic aneurysm. Psoriasis is associated with an increased risk of AA regardless of the disease severity (severe psoriasis: HR, 1.51, 95%CI, 1.04 to 2.19, *I*^2^ = 40.2% ; mild psoriasis:HR, 1.24, 95%CI, 1.08 to 1.42, *I*^2^ = 24.1%). However, the risk of AA was not significantly increased in female psoriasis patients (HR, 1.55, 95%CI, 0.65 to 3.72), patients with hypertension (HR, 1.44, 95%CI, 0.85 to 2.42), patients with hyperlipidemia (HR, 1.69, 95%CI, 1.15 to 2.48) and patients with diabetes (HR, 1.15, 95%CI 0.46 to 2.87).

**Table 2.**
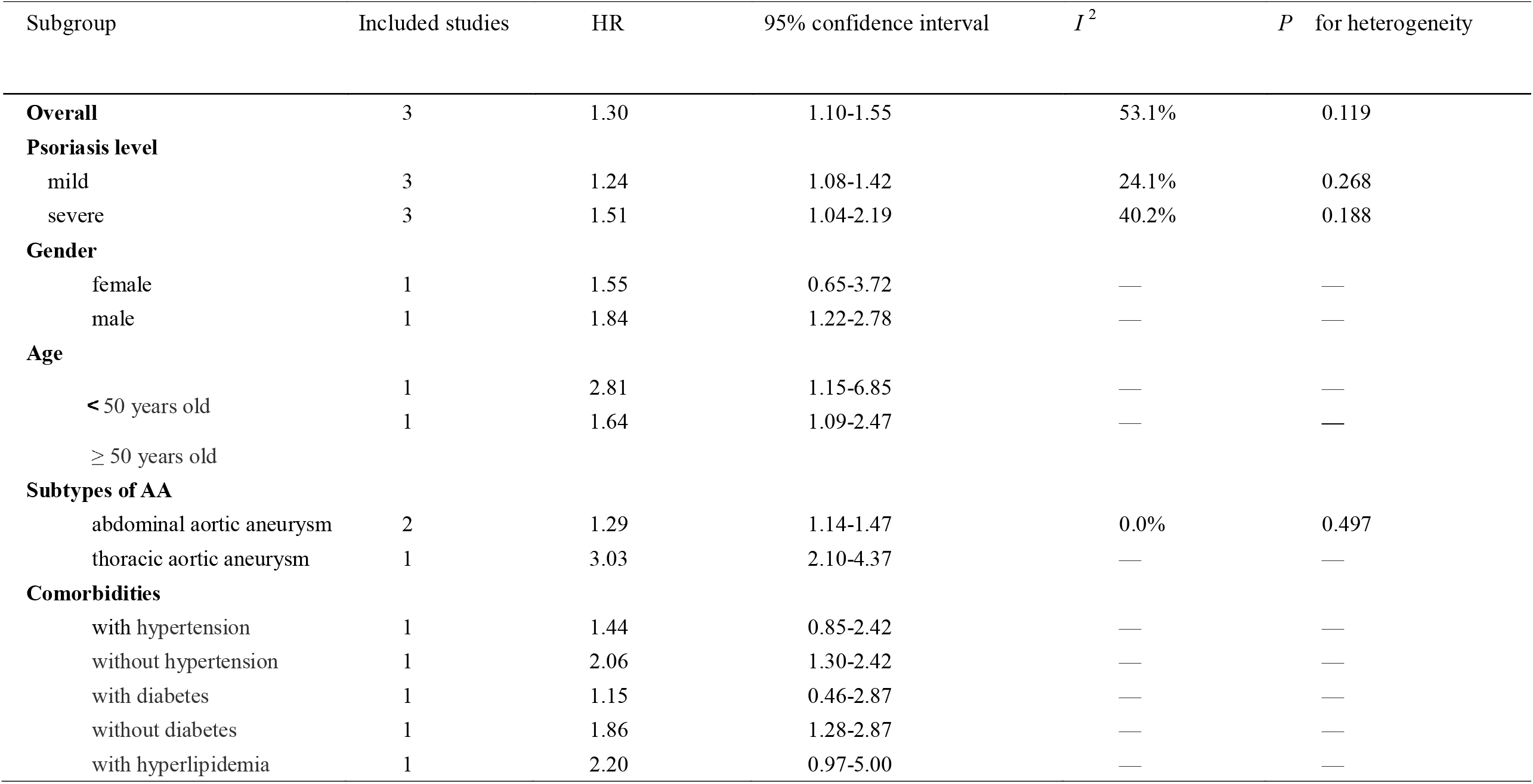
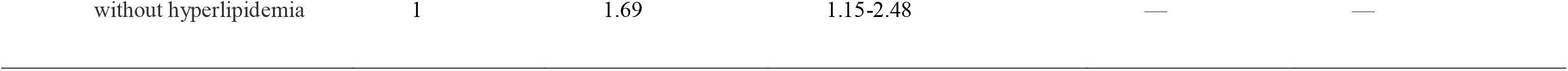
Subgroup analysis

## 4. Discussion

This is the first systematic review to estimate the risk of AA in patients with different degrees of psoriasis. It showed that psoriasis patients are more susceptible to aortic aneurysms compared to the general population regardless of disease severity of psoriasis. Moreover, the association was not significant in female psoriasis patients, patients with hypertension, patients with hyperlipidemia and patients with diabetes.

Definitions and inclusion criteria of exposure and outcome were slightly different between included studies (see Supplementary Table 2), and this led to inevitable clinical heterogeneity. Subgroup analyses showed gender and comorbidities (hypertension, hyperlipidemia, diabetes) affect risks of psoriasis. Three included studies tested the risk of AA in mild/severe psoriasis, but the definition of disease severity is different, and thus we failed to investigate the relationship between the risk of AA and disease severity of psoriasis. Diabetes is associated with a reduced incidence of AA[14, 15], and it may decrease the risk of AA in psoriasis patients. Some other factors (e.g. smoking, family history of AA) are strongly affect the risk of AA, but these were not tested in subgroup analyses by included studies, and thus we failed to include these factors in our results. Due to the limited study population and obvious heterogeneity, the statistical power of meta-analyses was strongly limited. Therefore, future large observational studies, however, to better eliminate confounders, are required on this field.

Psoriasis is an immune-mediated inflammatory disease associated with cardiometabolic comorbidities[5, 16]. Studies have confirmed psoriasis increases the risk of subclinical cardiovascular disease, as evidenced by higher coronary artery calcium[17] and an elevated burden of coronary artery disease[1, 18]. Elevated blood inflammatory biomarkers in patients with psoriasis indicate a moderate role of systemic inflammation in pathophysiology of psoriasis[19]. To identify concrete inflammatory lesions, researches detected metabolic activity in vessels and other tissues using ^18^FDG-PET/CT[1, 7, 20, 21, 22], and researchers observed significantly increased aortic inflammation in psoriasis patients. Currently, inflammation was considered to have a crucial pathogenic role in the development and progression of AA[8]. Psoriasis is an immune-mediated genetic disease[23] and AA could be an acquired disorder. Psoriasis may be associated with the risk of connective tissue disorders which could increase the risk of AA[24]. Therefore, natural course of psoriasis might promote the development of AA. Otherwise, hemodynamic factors and aortic stiffening could also contribute to AA development[10, 25]. In patients with psoriasis, increased arterial stiffness is presented[26] and it is associated with systemic inflammation[27]. Aortic stiffening leads to axial stress which then induces and augments processes necessary for AA growth such as inflammation and aortic wall remodeling[25].

From a preventive point of view, AA screening is strongly suggested recently[28]. In light of present findings, AA screening might be conducted among patients with psoriasis. Severe skin inflammation of psoriasis indicates elevated aortic inflammation[1], thus increasing the probability of the development of AA. AA screening of psoriasis with higher psoriasis area severity index score could be more effective and economical. From a therapeutic perspective, anti-inflammatory agents such as TNF-α antagonist [29, 30] could reduce vascular inflammation and improve endothelial function. Currently, no effective drug therapy is available for AA, and anti-inflammatory agents may become potential drugs limiting the development and progression of AA.

When interpreting results of this systemic review, several limitations should be considered. However, we failed to conduct sensitivity analyses and a meta-regression of confounds for lack of enough studies. Substantial clinical heterogeneity was presented in meta-analyses, and pooled analyses included no more than three studies, and thus results of this systematic review should be interpreted carefully.

## 5. Conclusion

Our findings indicate that psoriasis may be related with a higher incidence of AA. However, limited studies were conducted on this topic, and future epidemiological studies, however, to better eliminate confounders, are required on this field. Whether AA screening is effective and feasible in patients with psoriasis still require further researches.

## Data Availability

This article is a systematic review and meta-analysis, in which data were extracted from published studies.

## Acknowledgments

Authors would like to thank all the authors of the original articles.

## Ethical approval

Ethical approval was not necessary for this systematic review.

## Transparency declarations

None to declare

## Contributorship Statement

XY, XF and SC conceived the idea. Literature research was conducted by XY and XF. Study screening, study inclusion and quality assessment were conducted by XY and LX. Data extraction and analysis were carried out by XY and XF. XW and SC are guarantors. The manuscript was completed with the participation of all authors, and all authors approved the final manuscript.

## Figure legends

**Table 1.** Detailed characteristics of included studies

Abbreviations: AA, aortic aneurysm; ICD, international classification of diseases.

**Table 2.** Subgroup analyses

Abbreviations: AA, aortic aneurysm; HR, hazard ratio.

## Electronic Supplementary Material

**Supplementary Table 1**. Newcastle-Ottawa scale for assessing the methodological quality of included studies

**Supplementary Table 2**. Baseline characteristics of the study population

**PRISMA check list**

